# U.S. county-level characteristics to inform equitable COVID-19 response

**DOI:** 10.1101/2020.04.08.20058248

**Authors:** Taylor Chin, Rebecca Kahn, Ruoran Li, Jarvis T. Chen, Nancy Krieger, Caroline O. Buckee, Satchit Balsari, Mathew V. Kiang

## Abstract

**Background:** The spread of Coronavirus Disease 2019 (COVID-19) across the United States confirms that not all Americans are equally at risk of infection, severe disease, or mortality. A range of intersecting biological, demographic, and socioeconomic factors are likely to determine an individual’s susceptibility to COVID-19. These factors vary significantly across counties in the United States, and often reflect the structural inequities in our society. Recognizing this vast inter-county variation in risks will be critical to mounting an adequate response strategy.

**Methods and Findings:** Using publicly available county-specific data we identified key biological, demographic, and socioeconomic factors influencing susceptibility to COVID-19, guided by international experiences and consideration of epidemiological parameters of importance. We created bivariate county-level maps to summarize examples of key relationships across these categories, grouping age and poverty; comorbidities and lack of health insurance; proximity, density and bed capacity; and race and ethnicity, and premature death. We have also made available an interactive online tool that allows public health officials to query risk factors most relevant to their local context.

Our data demonstrate significant inter-county variation in key epidemiological risk factors, with a clustering of counties in certain states, which will result in an increased demand on their public health system. While the East and West coast cities are particularly vulnerable owing to their densities (and travel routes), a large number of counties in the Southeastern states have a high proportion of at-risk populations, with high levels of poverty, comorbidities, and premature death at baseline, and low levels of health insurance coverage.

The list of variables we have examined is by no means comprehensive, and several of them are interrelated and magnify underlying vulnerabilities. The online tool allows readers to explore additional combinations of risk factors, set categorical thresholds for each covariate, and filter counties above different population thresholds.

**Conclusion:** COVID-19 responses and decision making in the United States remain decentralized. Both the federal and state governments will benefit from recognizing high intra-state, inter-county variation in population risks and response capacity. Many of the factors that are likely to exacerbate the burden of COVID-19 and the demand on healthcare systems are the compounded result of long-standing structural inequalities in US society. Strategies to protect those in the most vulnerable counties will require urgent measures to better support communities’ attempts at social distancing and to accelerate cooperation across jurisdictions to supply personnel and equipment to counties that will experience high demand.

## Introduction

The spread of Coronavirus Disease 2019 (COVID-19) across the United States confirms that not all Americans are equally at risk of infection, severe disease, or mortality. Researchers have noted geographic disparities in critical medical resources, such as ventilators, hospital beds, and intensive care unit beds (1–4), which are important determinants of COVID-19 survival. Preliminary data from the epidemic in the US is exposing a convergence of demographic and socioeconomic issues that make low-income communities and people of color more vulnerable than others. Reports from Detroit, Milwaukee, New Orleans, and Chicago suggest that blacks comprise a disproportionate proportion of COVID-19 cases and deaths relative to their share of the population (5,6).

Age and pre-existing health conditions are recognized as important biological risk factors for serious disease and mortality from COVID-19 (7,8). Demographic factors including population density; living arrangements, such as group quarters (*e.g*., adult living facilities, nursing homes, or prisons), household size, household composition (*e.g*., grandparents living with grandchildren); or the number of educational institutions, for example, affect contact patterns and transmission rates at the county level (9–13). Socioeconomic factors like poverty, job insecurity, and lack of health insurance will determine the ability of populations to work-from-home and “shelter in place” (14–16), at a time when such non-pharmaceutical interventions are considered a primary defense against the outbreak (17,18).

Understanding the distribution of these county-specific characteristics is therefore critical to mounting an adequate, timely, and comprehensive response. Inter-county differences are particularly important to consider in the context of supportive local policies around social distancing as the epidemic unfolds, and for the relaxation of social distancing in the coming months. These structural issues must also be considered as we look ahead to possible scenarios for ending the epidemic, and to ways in which we can reconfigure health systems to be resilient to pandemic risks.

To assess each county’s level of risk of high COVID-19 medical burden, we examine available data for a range of characteristics for all U.S. counties, or county equivalents. We have made available an online tool for planners to explore the burden of these risk factors and others in their counties (Supplemental Materials Text S1).

## Methods

Using publicly available county-specific data from the Area Health Resources Files (19), American Community Survey (20), Centers for Disease Control and Prevention Atlas file (21), National Center for Health Statistics (22), and RWJF Community Health Rankings (23), we identified key biological, demographic, and socioeconomic characteristics influencing susceptibility to COVID-19, guided by international experiences, and consideration of epidemiological parameters of importance. We created five bivariate county-level maps to illustrate examples of key relationships across these categories grouping age and poverty; comorbidities and lack of health insurance; proximity, density and bed capacity; and race and ethnicity, and premature mortality. Bivariate maps show the risk in a county across independent axes, highlighting both the convergence and divergence of factors.

We have purposefully avoided weighting these risk factors relative to each other, since there is no strong evidence to rigorously assign importance across categories. The included univariate maps demonstrate the vast heterogeneity in risks across the United States (Supplemental Materials). The bivariate maps may assist county-and state-level policymakers in their response to COVID-19 by exemplifying how different risk factors together contribute to counties’ overall level of COVID-19 burden.

An online dashboard (Supplemental Materials Text S1) allows querying of these risk factors to create other bivariate maps that may be more informative to a given locality. The dashboard also includes an option to show counties above different population thresholds. Moreover, the appropriate covariate threshold for any state or county will depend on the context of that count; therefore, the online dashboard allows interested readers to change the threshold for each covariate. All our data and code are publicly available so they may be used to create more nuanced analysis, inform existing models, and shape policy. (Supplemental Materials Text S1).

## Results

### Age and poverty

Age (Figure S1) is an important biological risk factor for severe disease and mortality from COVID-19. In a study of 44,672 confirmed cases of COVID-19 from China, the case-fatality rate was highest among the elderly (7). In the United States, among 2,449 cases from February 12 to March 16, 2020, 53% of ICU admissions and 80% of deaths occurred among adults aged ≥ 65, with the highest percentage of severe outcomes among adults aged ≥ 85 years (8). Poverty (Figure S2), independently, heightens vulnerability to COVID-19, due to its association with higher risk of comorbidities, decreased access to care (15,16,24), and reduced ability to practice social distancing (25). During the H1N1 2009 pandemic, age-adjusted rates of hospitalization were found to be significantly higher in high-poverty neighborhoods relative to low-poverty neighborhoods in New York City (26). We are already beginning to see the differential effects of COVID-19 across communities, with higher burden found in high poverty areas in the United States (27,28).

Overall, 10% of the US population is over the age of 70 (29), and 12% of the population lives below the poverty line,(30). Figure 1 shows the distributions of age and poverty by county, highlighting the convergence of risk factors in southern states in particular. In 768 of 3,111 counties, at least 15% of the population is over 70 years old. In 667 of 3,106 counties, more than 20% of households live below the poverty line. 111 of 3,106 counties, accounting for about 1.6 million people, have both a high number of households below the poverty line and a high proportion of elderly residents. Georgia, Texas, and Arkansas are the states with the largest number of such counties. We discuss the compounding effect of poverty on age in subsequent sections.

**Figure 1.**
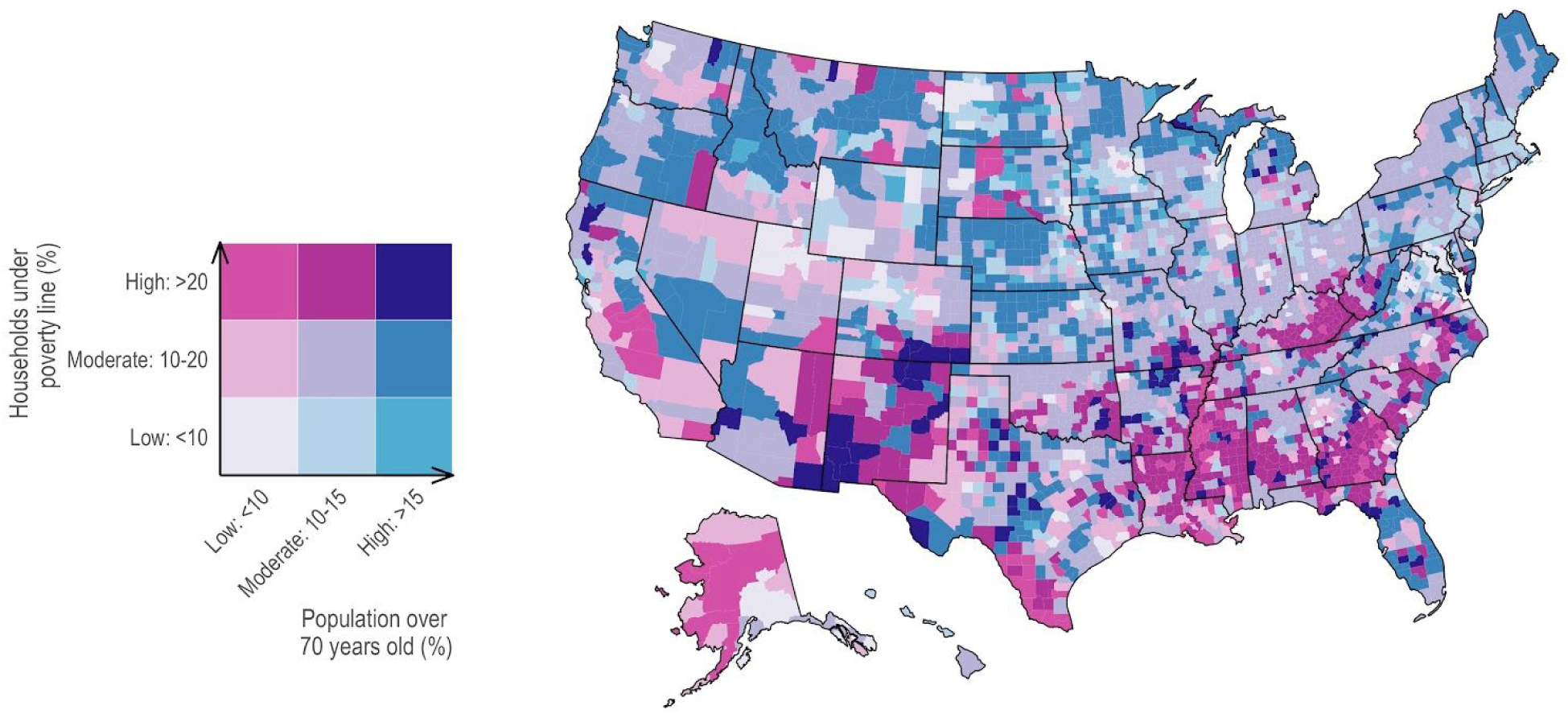
Percentage of households living in poverty, 2016 (Source: CDC Atlas via the Census Small Area Income and Policy Estimates) and percentage of population 70 years or older, 2018 (Source: National Center for Health Statistics Bridged Race Population Estimates 2018, Vintage 2018). Counties in the Southern US have high levels of poverty (dark reds). Counties in the Midwest have lower rates of poverty but high proportions of older residents (dark blues). 111 counties have both high levels of poverty and an aging population (dark purple). States like Texas exhibit high inter-county variability in risks.

### Comorbidities and lack of health insurance

Pre-existing health conditions, including hypertension, diabetes, and coronary heart disease, increase the likelihood of severe disease and mortality from COVID-19. In a study from China, the overall case-fatality rate was estimated at 2.4%, but was markedly higher for those with cardiovascular disease (10.5%), diabetes (7.3%), chronic respiratory disease (6.3%), and hypertension (6.0%) (7). The prevalence of non-communicable diseases varies across the US. The 55 of 3,106 counties, where at least 20% of the population over 20 years old carries a diagnosis of diabetes, are primarily concentrated in the Southeast US, with the most in Georgia, Texas, and Alabama, consistent with previous research (Figure S3) (31,32).

Individuals without health insurance (Figure S4) have limited access to hospital care (33). Those without health insurance are also more likely to work in service-oriented industries with decreased ability to work from home (34), putting them at higher risk of infection. Within weeks of implementing social distancing measures, and closures of schools and non-essential businesses, unemployment in the United States rose from 282,000 to 3.3 million (35). The consequence of these unemployment rates on health insurance are not captured in our data.

We examine hospitalization rates of coronary heart disease (CHD) and hypertension (HTN) among Medicare beneficiaries, and the health insurance status of adults 18-64 years old (Figure 2). In addition to increasing the burden of severe disease and mortality from COVID-19 in a county, high baseline rates of hospitalization for CHD and HTN among the elderly affect availability of hospital and ICU beds. Populations with high proportions of uninsured 18-64 year olds are likely to be in work or living situations that increase their risk of exposure and transmission. Together these variables inform county-level risks across a range of age groups and represent the overall level of COVID-19 burden in a county. In 661 of 3,099 counties, the hospitalization rates for patients with CHD and HTN was >250 per 1,000 Medicare beneficiaries (2014-2016), compared to the national average of 200. In 180 of 3,111 counties, at least 25% of the population aged 18-64 years lacks health insurance. Approximately 2.2 million residents live in the 40 counties that fall in both high categories, and 26 of these counties are in Texas.

**Figure 2.**
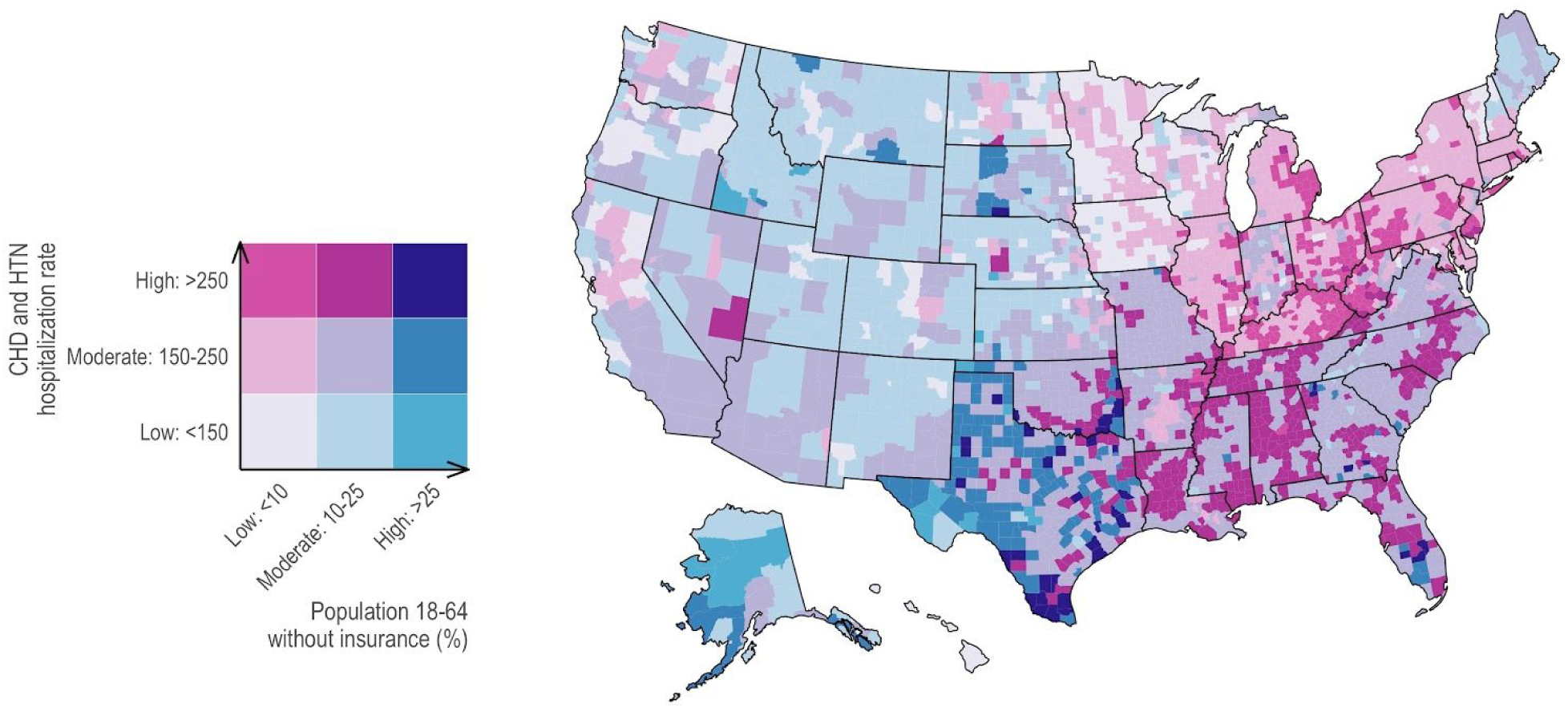
Coronary heart disease and hypertension hospitalizations per 1,000 Medicare beneficiaries (65+), 2014-2016 (Source: CDC Atlas via the Centers for Medicare and Medicaid Services Medicare Provider Analysis and Review file) and percentage of 18-64 year olds with no health insurance, 2017 (Source: Area Health Resources Files via the Census Small Area Health Insurance Estimates). Counties in the South have high rates of hospitalizations and moderate proportions of populations without health insurance (dark reds). Counties in the West have lower rates of hospitalizations and lower proportions of people without insurance (light blues). 40 counties, two-thirds of which are in Texas, have both high CHD and hypertension hospitalization rates among the Medicare beneficiaries, and a high proportion of the population aged 16-64 without insurance (dark purple).

### Proximity, density and bed capacity

Household size and characteristics like the number of generations living together (Figure S5), will affect transmission rates, household vulnerability, and the feasibility of social distancing measures. A study looking at transmission among close contacts in Shenzhen, China estimated a secondary attack rate of approximately 15% among household contacts (36). The prevalence of group quarters, such as correctional facilities, nursing homes, student housing, and homeless shelters, among others (37), also increases the risk of transmission for members of these communities. 527 of 3,111 counties have greater than 5% of their population living in group quarters (Figure S6).

Additionally, high population density increases risk of rapid transmission, especially in the absence of adequate or timely social distancing measures, as evidenced by the current scale of the outbreak in New York City (38). 425 of 3,111 counties, mostly concentrated in the eastern United States and on the West coast, have greater than 100 people living per square mile, compared to the national median of 45 people per square mile (Figure S7).

The ability of counties to respond to these additional risks will partly depend on local bed capacity. Several studies examining hospital capacity in the U.S. predict substantial shortages in hospital-and ICU-beds (1–4). 949 of 3,111 counties have fewer than 100 hospital beds per 100,000 people, as compared to the national median of 185 (Figure S8). 1,939 of 3,111 counties have fewer than 10 ICU beds per 100,000 population (Figure S9).

In Figure 3 we examine the overlapping risks of group quarters, population density, and hospital bed capacity. 371 of 3,111 counties, with over 10 million residents, have both a substantial proportion of the population living in group quarters and low ICU bed capacity (Figure 3A), putting them at heightened risk of facing critical care capacity shortages. 155 of 3,111 counties across the United States are both densely populated and have low hospital bed capacity per capita, and are home to over 17 million people (Figure 3B). More than 40% of these counties are concentrated in Georgia, Virginia, Ohio, Kentucky, and Michigan, and are at higher risk of being overwhelmed in the event of an outbreak (Figure 3B).

**Figure 3A.**
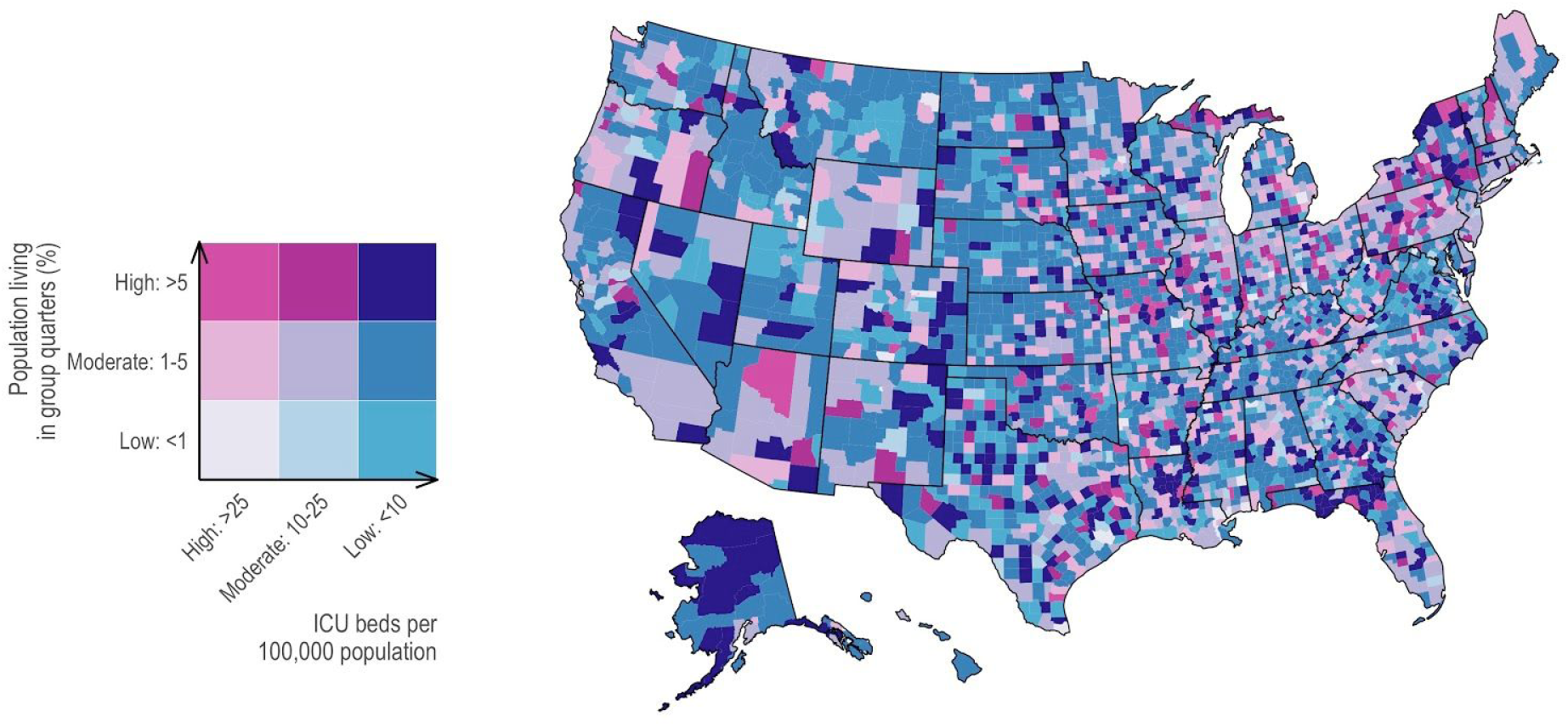
Percentage of population living in group quarters, 2018 (Source: American Community Survey) and intensive care unit (ICU) beds per 100,000 population, 2017. Intensive care unit beds are defined as medical/surgical, cardiac, and other intensive care beds (Source: Area Health Resources Files). There is high heterogeneity across the US: some counties have a substantial proportion of their population living in group quarters (dark reds), others with low rates of ICU beds (dark blues) and some with high levels of both (dark purple).

**Figure 3B.**
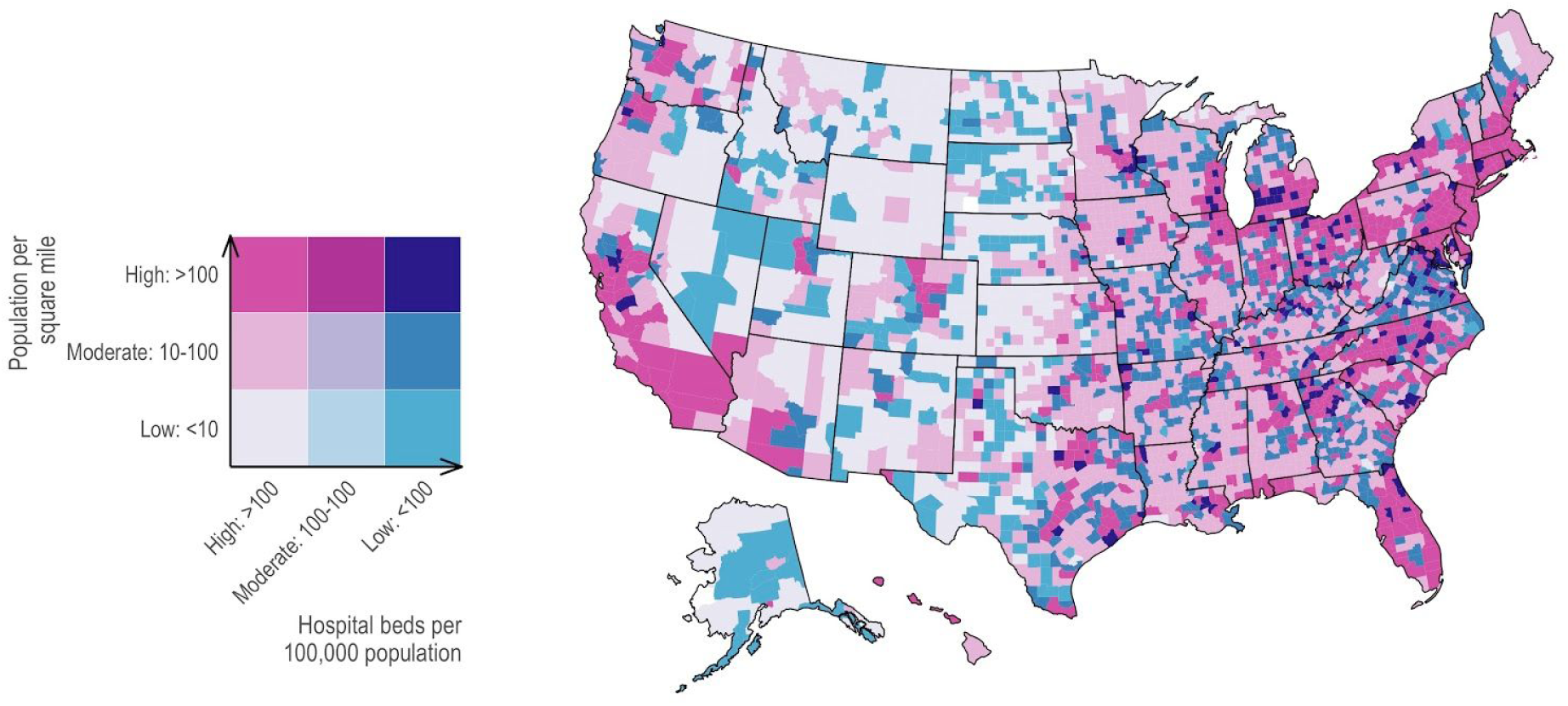
Population per square mile of land mass (Source: US Census Bureau) and hospital beds per 100,000 population (Source: Area Health Resources Files). Counties on the East and West coasts have high population density (dark reds), while rural counties have fewer hospital beds per 100,000 population, focused heavily (dark blues). Some counties have high levels of both (dark purple).

### Race and ethnicity and premature death (YPLL)

Persistent structural inequities in American society have resulted in health disparities that are closely associated with race and ethnicity in the United States (39). Communities of color do not only have increased risk of chronic diseases (40), which increase risk of COVID-19 infection, but also experience unequal access to health care (5,41). Populations of color are also disproportionately unemployed (42), incarcerated (43), and without health insurance (14) – all independent risks, as we have discussed above. These disparities contribute to higher age-adjusted death rates among non-Hispanic blacks relative to non-Hispanic whites (44), Even within levels of chronological age, groups subjected to economic and social deprivation have a higher risk of comorbidities and premature mortality compared to people who are chronologically older but more privileged (45,46).

Figure 4 shows the relationship between the proportion of the population that is non-Hispanic and non-White and age-adjusted years of potential life lost (YPLL) before age 75 per 100,000 population in 2017. In 366 of 3,111 counties, over 50% of the population is non-Hispanic and non-White. These communities are predominantly located in Southern states, in Texas, Georgia, and Mississippi. In 758 of 3,051 counties, there are at least 10,000 years of potential life lost (YPLL) before age 75 per 100,000 population in 2017, in comparison to the national median of approximately 8,100. 162 of 3,051 counties, with over 5.6 million residents, have a high proportion of the population that is non-Hispanic and non-White, and has a high number of years of potential life lost. Approximately 44% of these counties fall in the states of Mississippi, Georgia, New Mexico, and Alabama.

**Figure 4.**
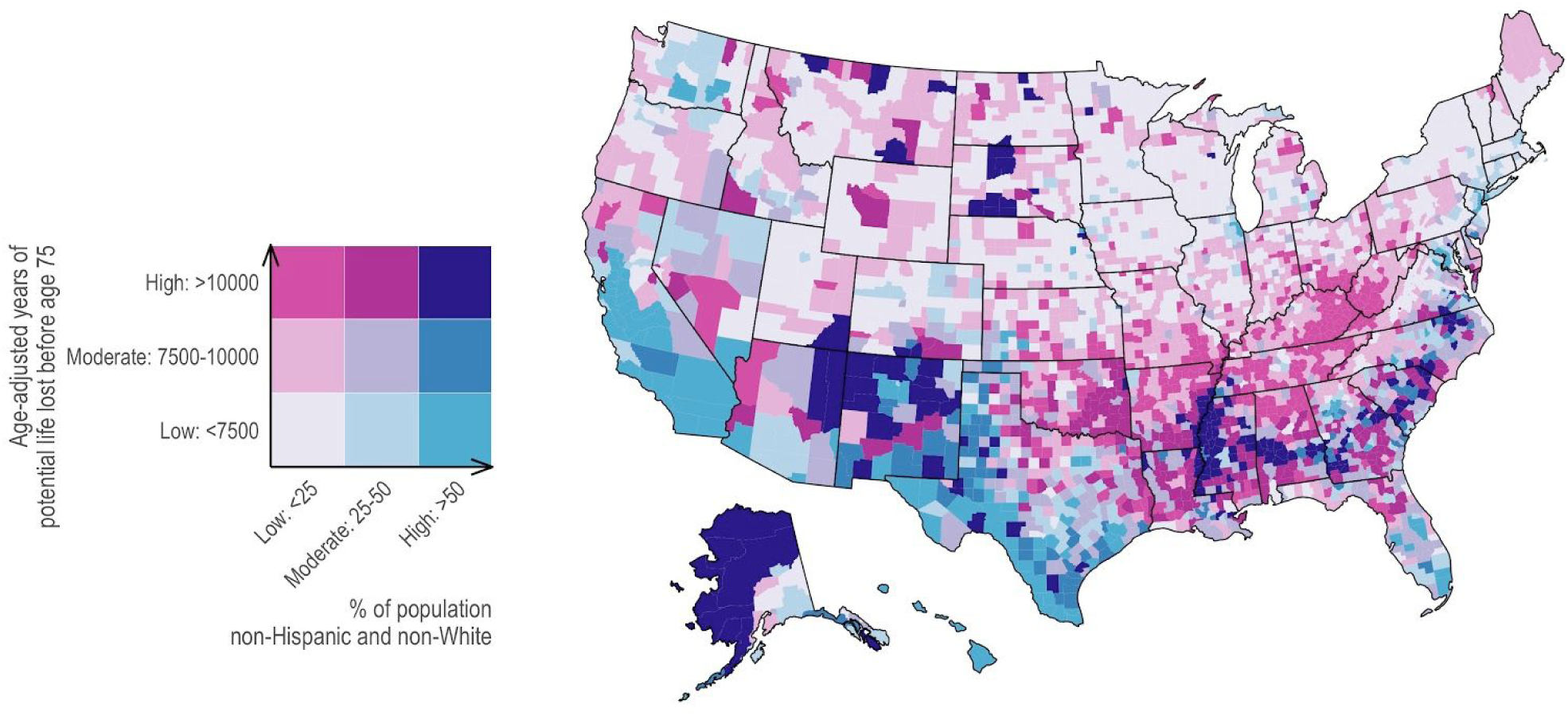
Age-adjusted years of potential life lost (YPLL) before age 75 per 100,000 population, 2017 (Source: Robert Wood Johnson Foundation County Health Rankings) and percentage of population non-Hispanic and non-White, 2018 (Source: National Center for Health Statistics). Counties with either high rates of YPLL before age 75 per 1000,000 population (dark reds), high proportion of non-Hispanic and non-White (dark blues), or high levels of both (dark purple) are largely concentrated in Southeastern and Southwestern counties.

## Discussion

Our findings demonstrate significant inter-county variation in the multiple, interconnected social determinants that will affect both the burden of COVID-19 disease and the ability of communities to effectively respond to it. More than 20% of households in nearly one in five counties in the United States are living below the poverty line. In more than 10% of all counties, more than half the population is non-Hispanic, non-White. Approximately 25% of all counties have a high proportion of elderly persons. Together, these counties represent millions of Americans at heightened risk of COVID-19 infection and risk of severe disease and death given infection.

While the East and West coast cities are particularly vulnerable due to their densities (and travel routes), a large number of counties in the Southeastern states have a multitude of interconnected risk factors, many of which can be traced back to legacies of Jim Crow and race relations in the South (47,48). Many Americans with chronic comorbidities, lack (and recent loss) of health insurance, inability to work from home, and poor access to care, are likely to be disproportionately affected by COVID-19, due to their increased risk of both infection and severe disease. The majority of counties that carry these multiple burdens, however, are also located in states that have been tepid in their social-distancing response (49). Our analysis provides insights into how communities of color may likely continue to bear disproportionately high burden of infection, severe disease, and mortality (5,50). Collecting and sharing data on COVID-19 outcomes by race/ethnicity, which surveillance systems hitherto have not been systematically reporting (51,52), will be crucial to understanding and rectifying inequities in the distribution of COVID-19 outcomes.

Attempts to avert the high burden among these populations will require a concerted strategy to ameliorate the conditions that compound their vulnerability, beyond merely ramping up bed capacity. These counties and states, in particular, will benefit from implementing measures that make it possible for individuals and families to practice social distancing, for example, through financial aid and expanded healthcare. These measures are especially crucial given that nearly 10 million Americans in the last two weeks of March have filed for unemployment (53).

In terms of medical response capacity, approximately 60% of all counties have fewer than 10 ICU beds per 100,000 population and 30% of counties have just over half the number of hospital beds per capita than the national median. In anticipation of heightened demands on healthcare systems in counties that are multiply vulnerable, states and the federal government need to ramp up inter-jurisdictional coordination efforts to move supplies and personnel to meet rapidly shifting local demands.

The risk factors described here are by no means a comprehensive list. Additional risk factors on the county-level, such as the proportion of workers in industries that preclude working remotely, language, immigration status, numbers of incarcerated and homeless persons, measures of inequality like the Gini coefficient and Index of Concentration at the Extremes (ICE), and density of residential drug treatment programs and residential mental health facilities, may all contribute to how counties are affected and respond. There currently is insufficient evidence to justify assigning weights to different risk factors, but as more data become available, future research may expand on our analysis by constructing a polysocial risk score (54).

## Conclusion

At a time when the US continues to inadequately test its population or protect healthcare providers, the need for actionable, contextually relevant data that allows for judicious distribution of our limited resources becomes imperative. County, state, and national planners will benefit from examining and preparing for the local factors that are likely to influence their counties’ ability to respond. By April 6, there were more than 330,000 cases in the United States, across all states, Washington D.C., and four U.S. territories (55). The morbidity and mortality toll, as early numbers indicate, will likely be disproportionately borne by communities of color in the absence of concerted, aggressive, and proactive responses across administrative boundaries that account for the vast inter-county variability in risks across populations.

## Data Availability

We provide data and code to reproduce this paper.

https://github.com/mkiang/county_preparedness/

## Acknowledgements

We thank Ayesha S. Mahmud, Nishant Kishore, and Tori Cowger for their valuable input and feedback.

## Supplemental Materials

<u>Supplemental Text S1. Interactive results viewer with reproducible code and data.</u> For interested readers, we provide an interactive results viewer at https://mkiang.shinyapps.io/county_risks/ which allows readers to select any combination of covariates discussed in this paper as well as a range of thresholds. We also provide data and code to reproduce this paper at https://github.com/mkiang/county_preparedness/.

## List of Figures

**Figure S1.**
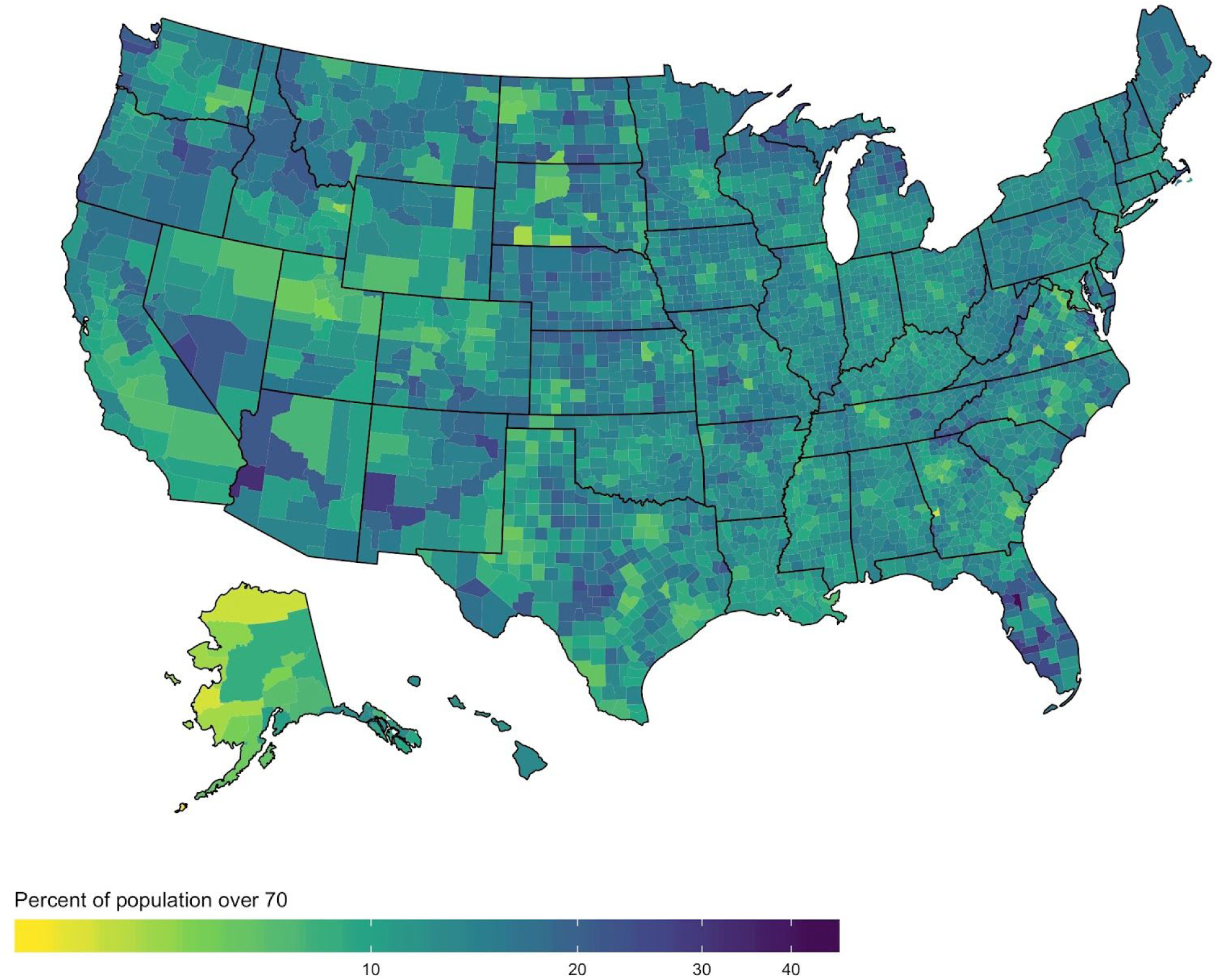
Percentage of population 70 years or older, 2018. Source: National Center for Health Statistics Bridged Race Population Estimates 2018, Vintage 2018.

**Figure S2.**
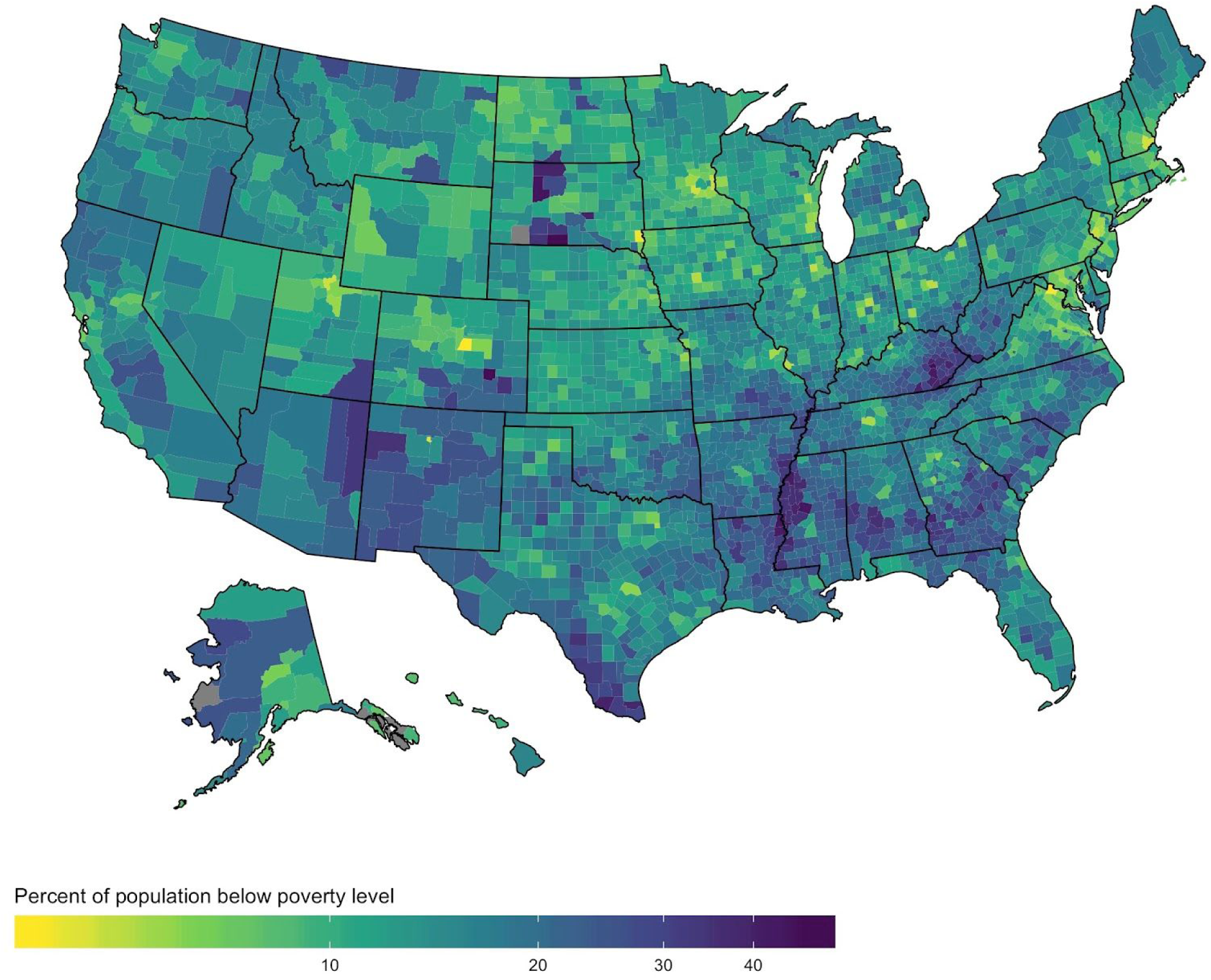
Percentage of households living in poverty, 2016. Source: CDC Atlas via the Census Small Area Income and Policy Estimates.

**Figure S3.**
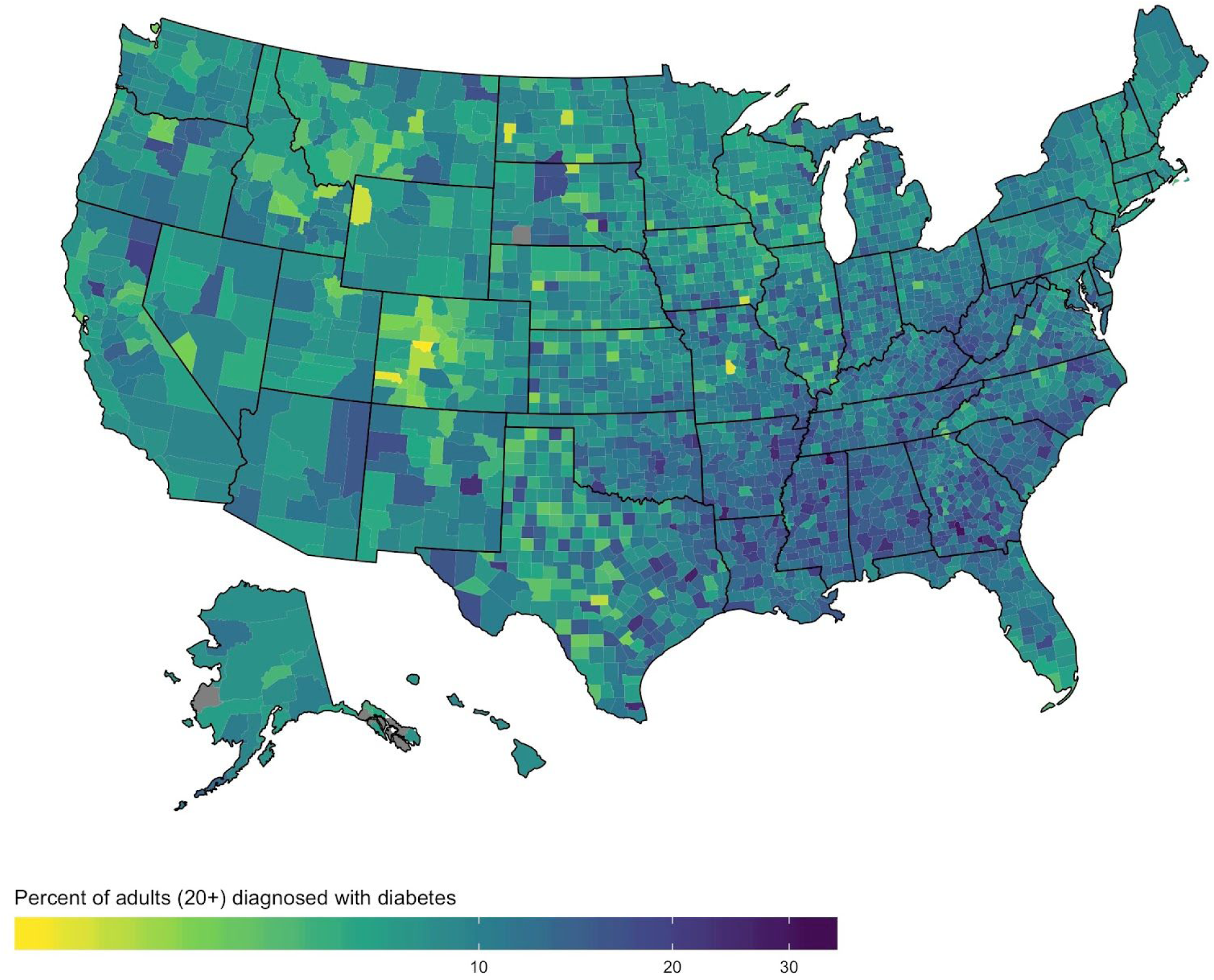
Age-adjusted percentage of adults over 20 years old who are diagnosed with diabetes, 2016. Source: CDC Atlas via the Behavioral Risk Factor Surveillance System.

**Figure S4.**
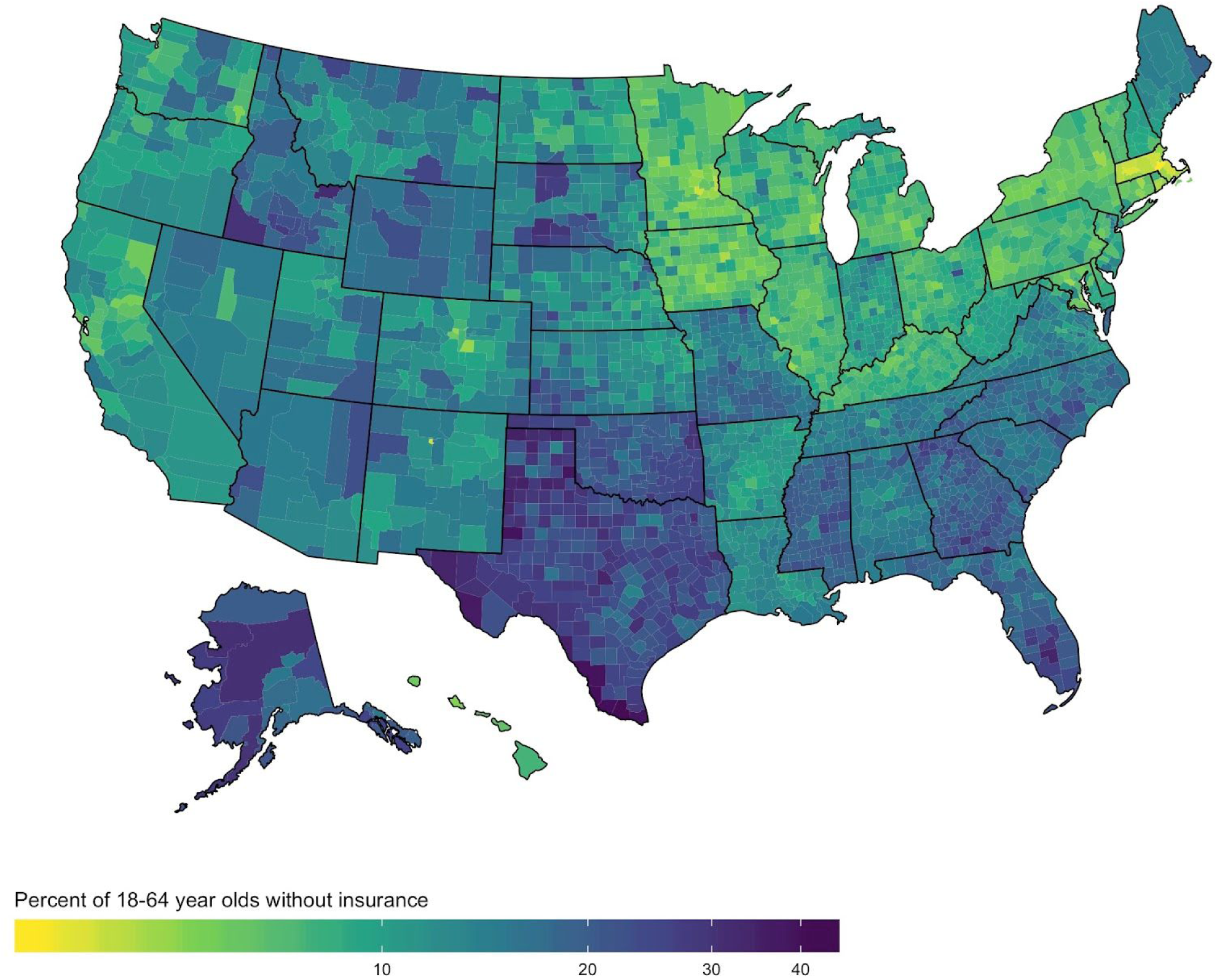
Percentage of 18-64 year olds with no health insurance, 2017. Source: Area Health Resources Files via the Census Small Area Health Insurance Estimates (variable F15498-17).

**Figure S5.**
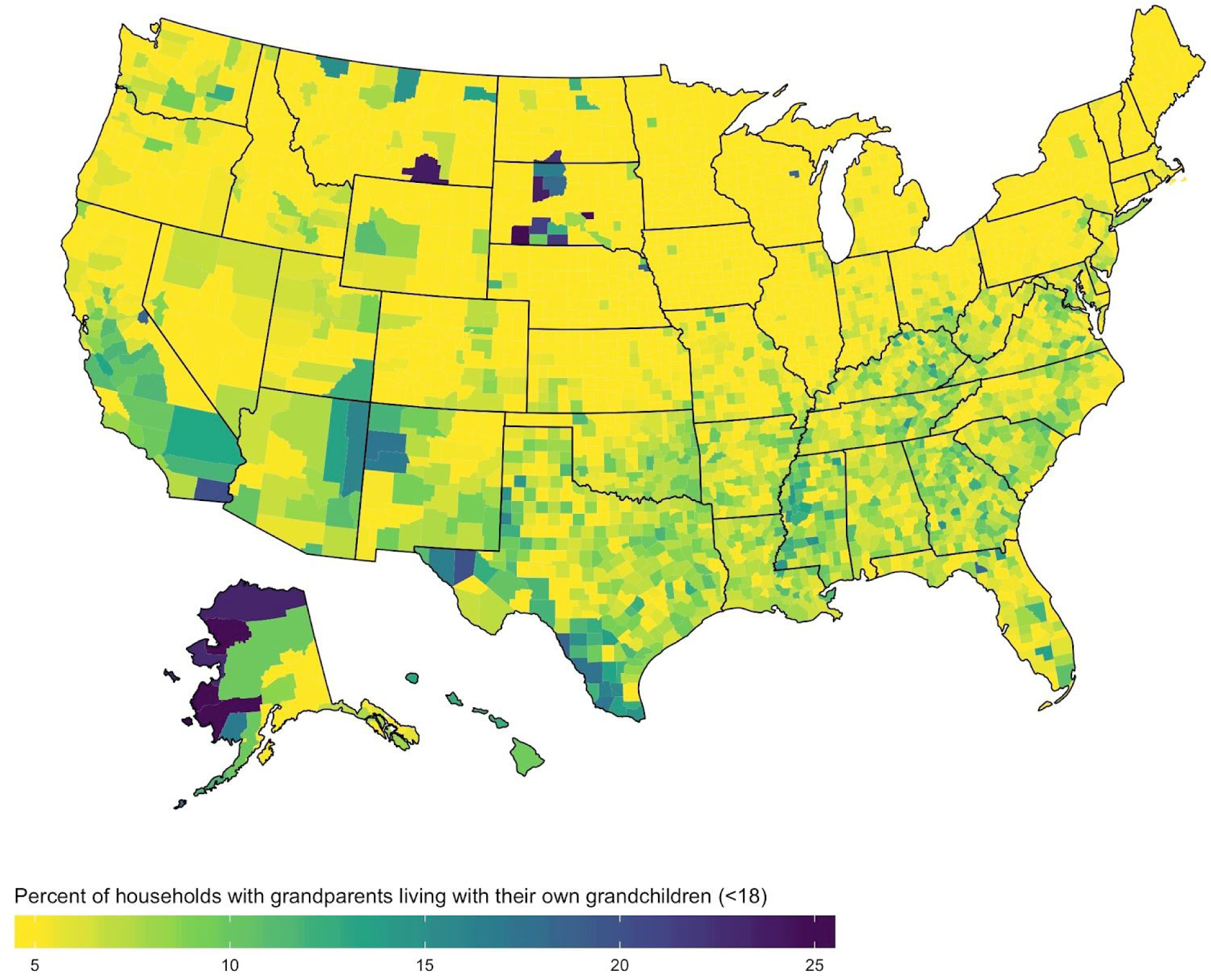
Percentage of households with grandparents living with their own grandchildren (<18 years old), 2018. Source: US Census Bureau’s American Community Survey 5-year, 2018 (variable DP02_0043PE).

**Figure S6.**
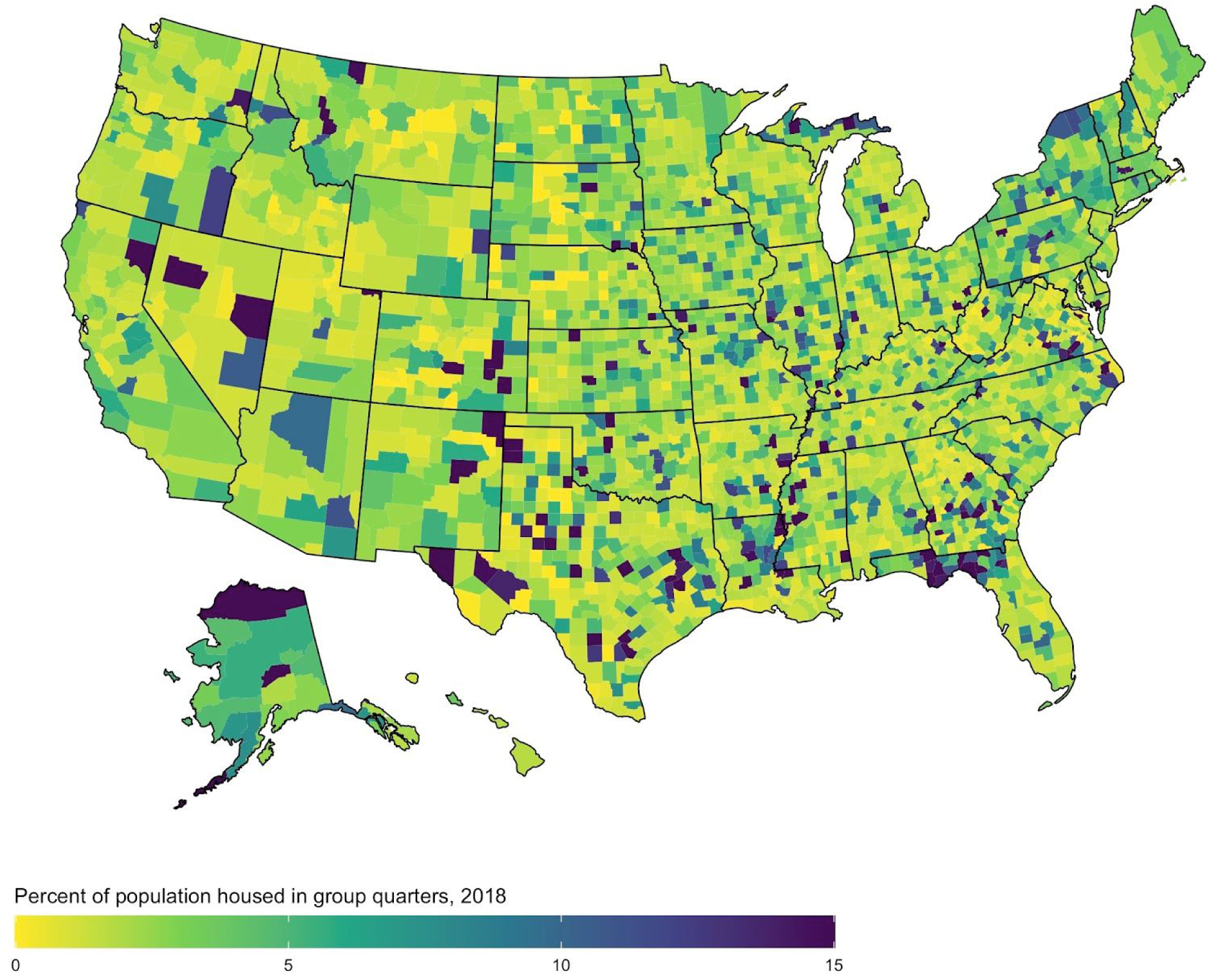
Percentage of population living in group quarters, 2018. Group quarters are defined by the US Census Bureau and include both institutional group quarters such as correctional facilities, nursing homes, or mental hospitals, as well as non-institutional facilities such as military bases, group homes, college dormitories, or shelters Source: US Census Bureau’s American Community Survey 5-year, 2018 (variable S1101_C01_020E).

**Figure S7.**
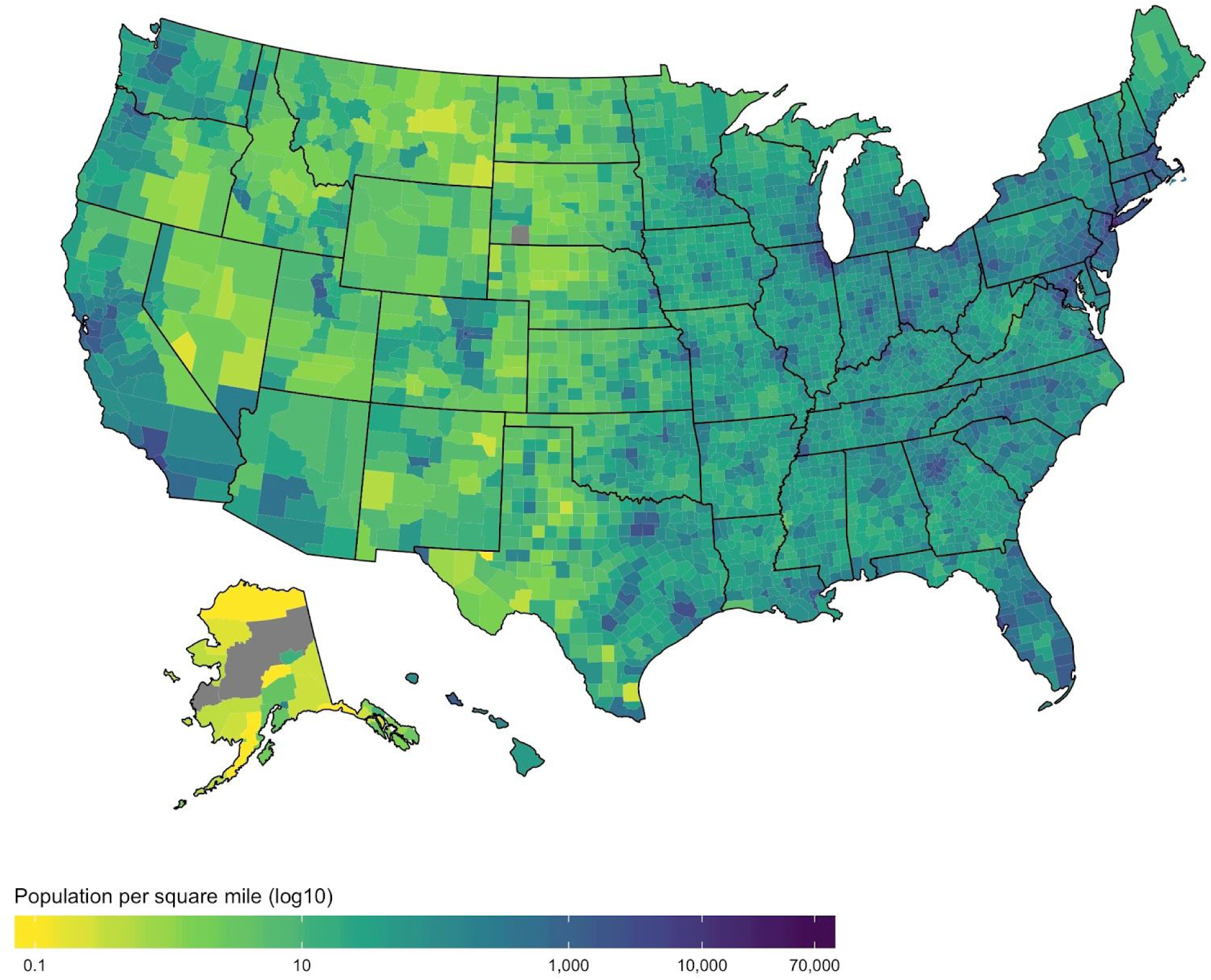
Population per square mile of land mass (log10), 2010. Source: US Census Bureau (summary table GCT-PH1).

**Figure S8.**
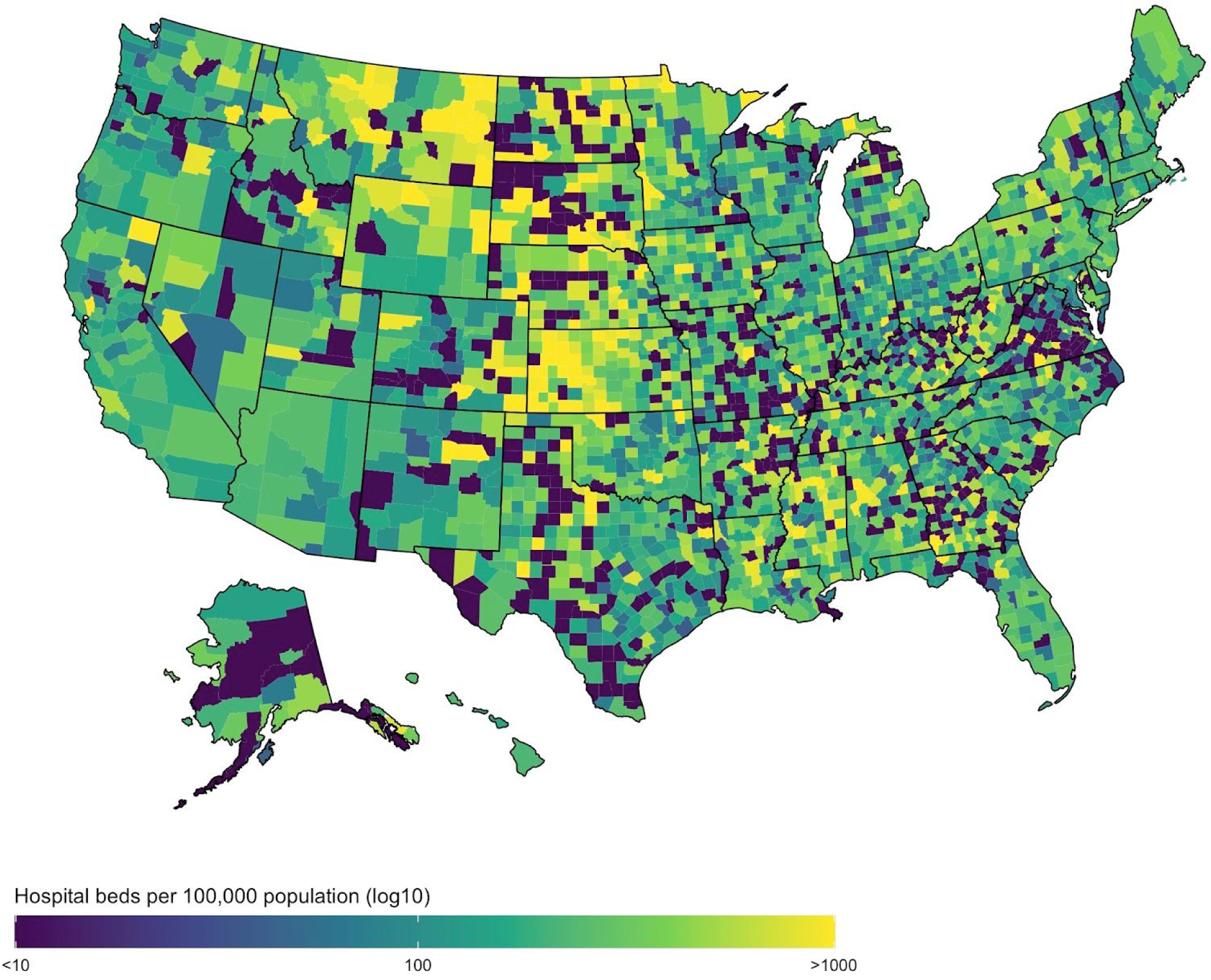
Hospital beds per 100,000 population, 2017. Source: Area Health Resources Files (variable F08921-17).

**Figure S9.**
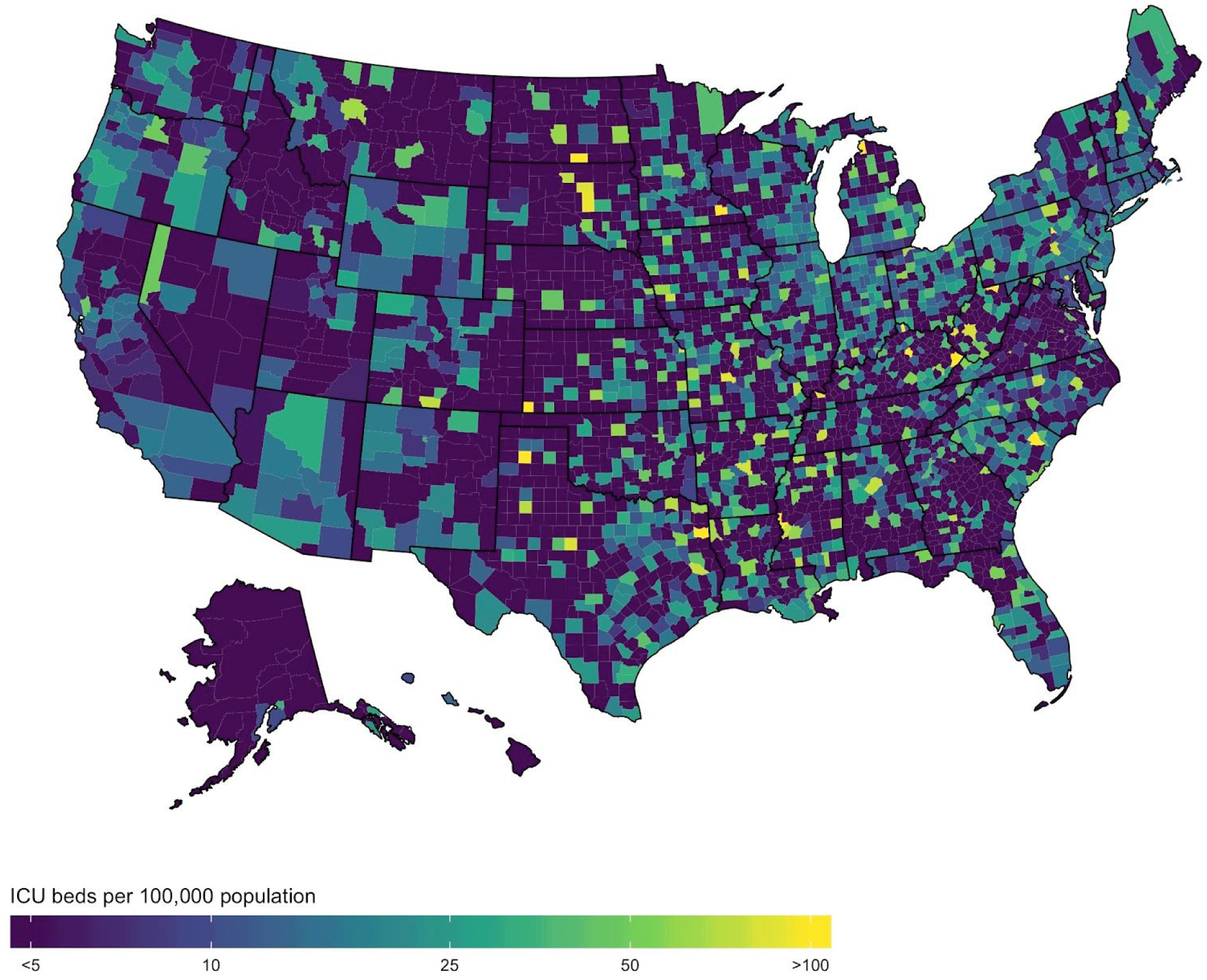
Intensive care unit (ICU) beds per 100,000 population, 2017. Intensive care unit beds are defined as medical/surgical, cardiac, and other intensive care beds. We exclude pediatric, neonatal, and burn intensive care beds. Source: Area Health Resources Files (variables F09139-17, F09133-17, F13309-17).

**Figure S10.**
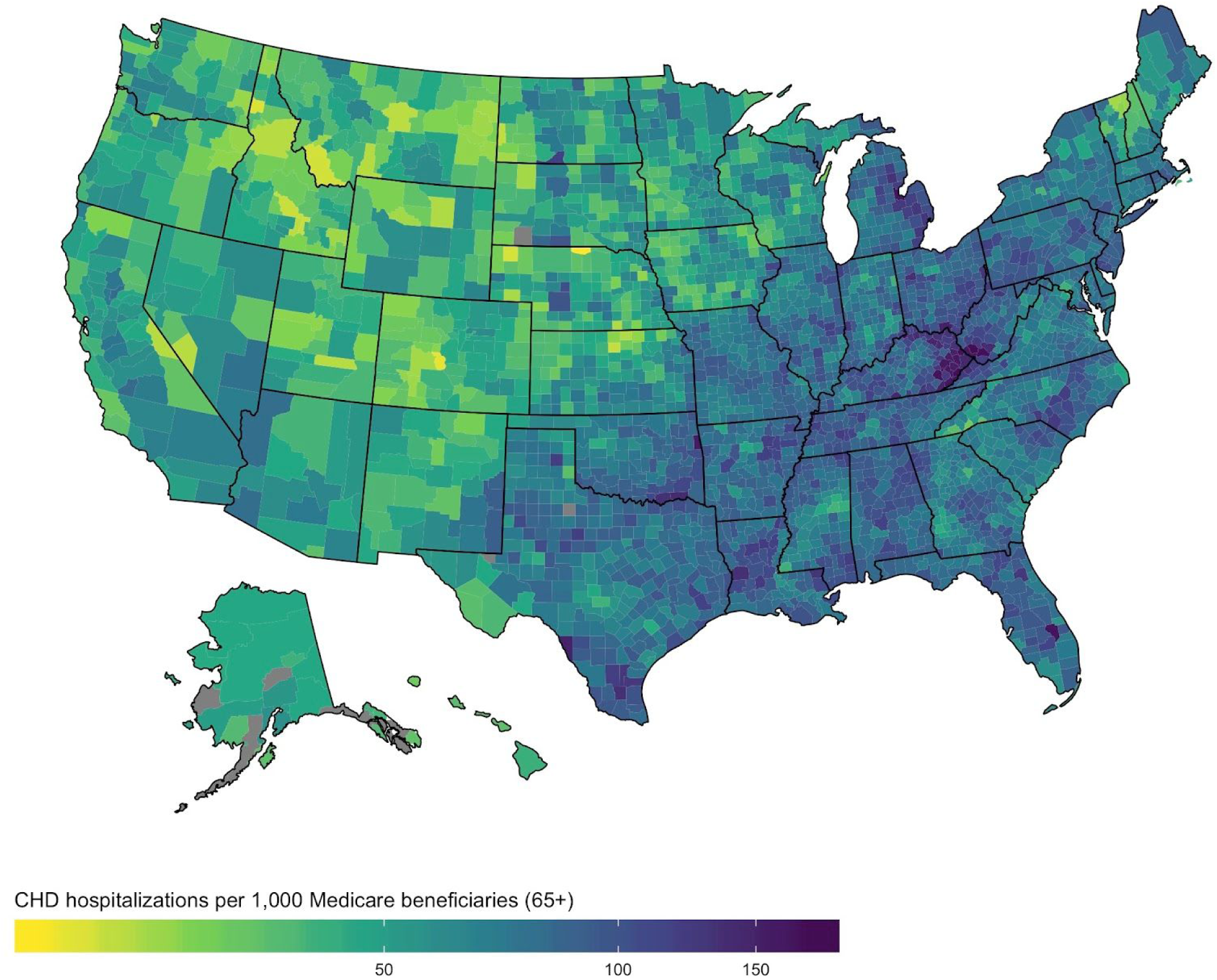
Coronary heart disease-related hospitalizations per 1,000 Medicare beneficiaries (65+), 2014-2016. Source: CDC Atlas via the Centers for Medicare and Medicaid Services Medicare Provider Analysis and Review file.

**Figure S11.**
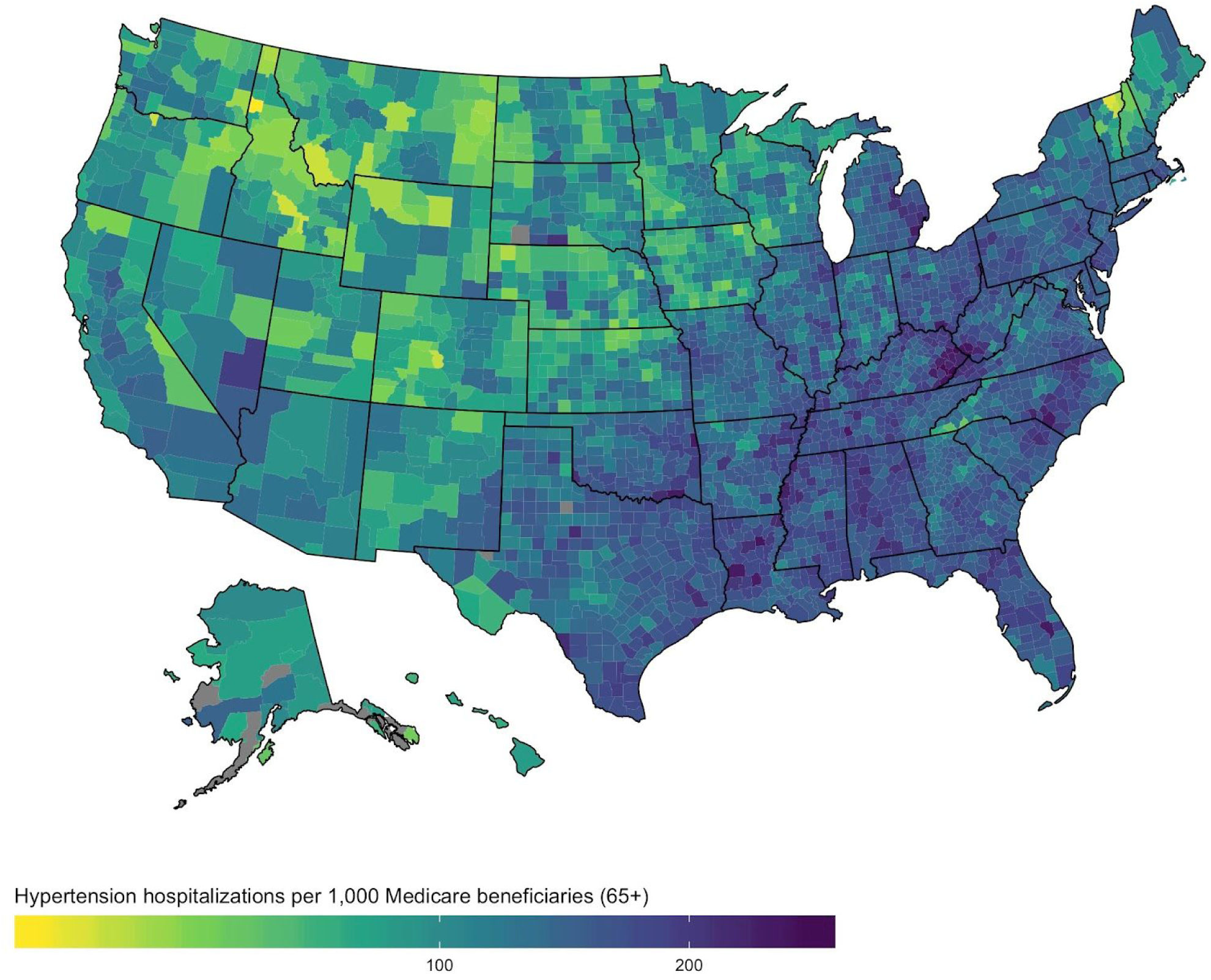
Hypertension hospitalization rate per 1,000 Medicare beneficiaries (65+), 2014-2016. Source: CDC Atlas via the Centers for Medicare and Medicaid Services Medicare Provider Analysis and Review (MEDPAR) file.

**Figure S12.**
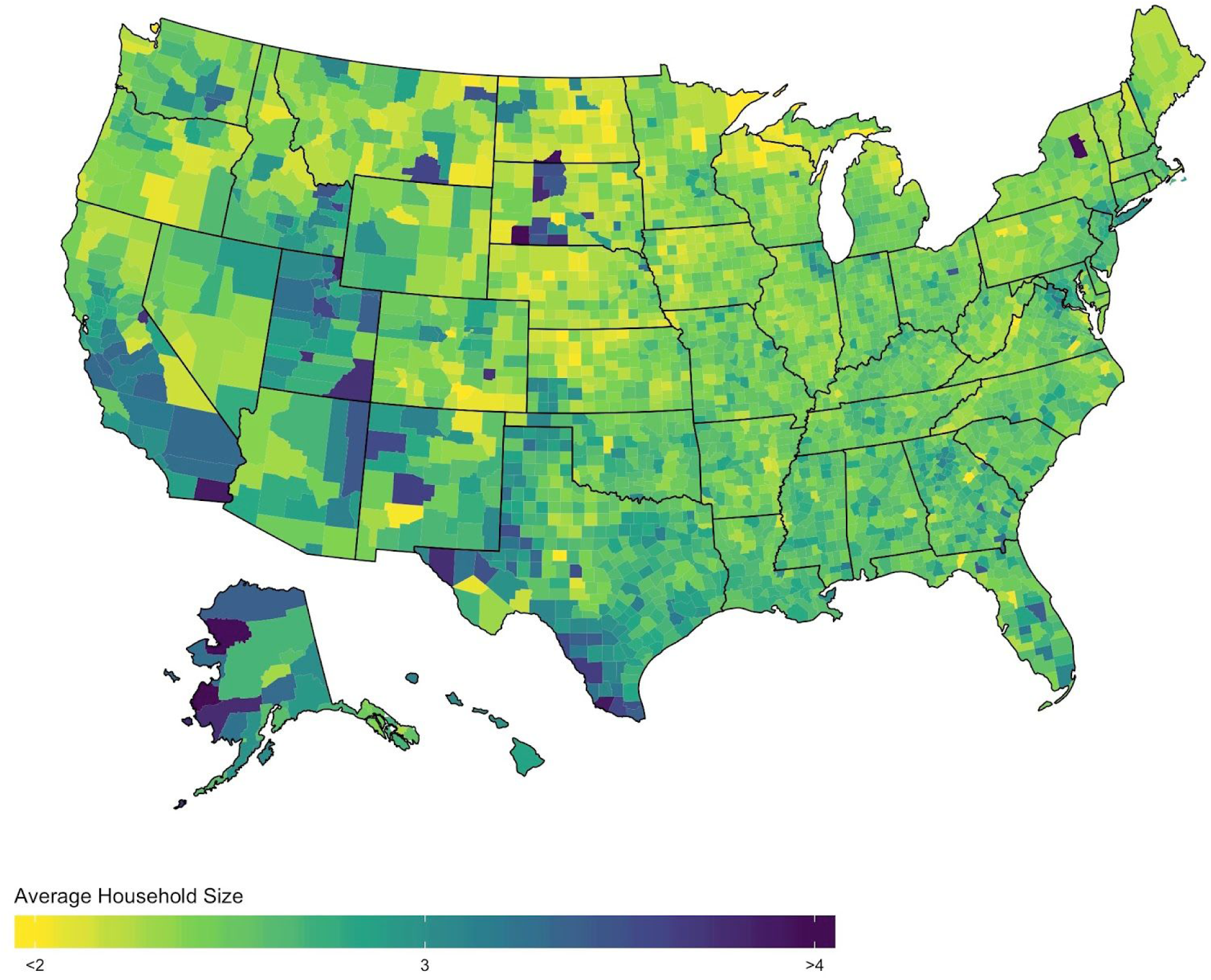
Average household size, 2018. Source: US Census Bureau’s American Community Survey 5-year, 2018 (variable DP02_0015E).

**Figure S13.**
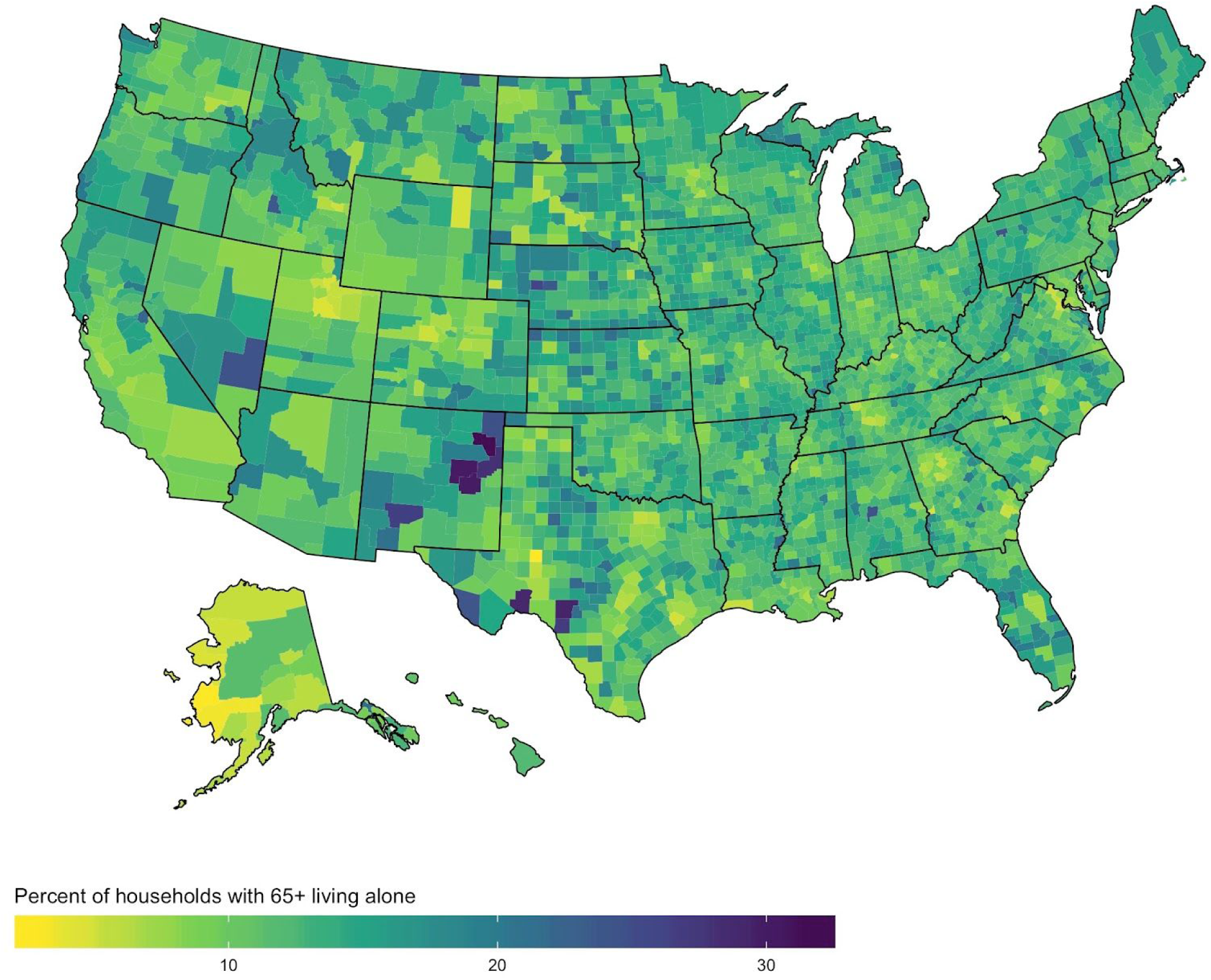
Percentage of households with 65+ year olds living alone, 2018. Source: US Census Bureau’s American Community Survey 5-year, 2018 (variable DP02_0012PE).

**Figure S14.**
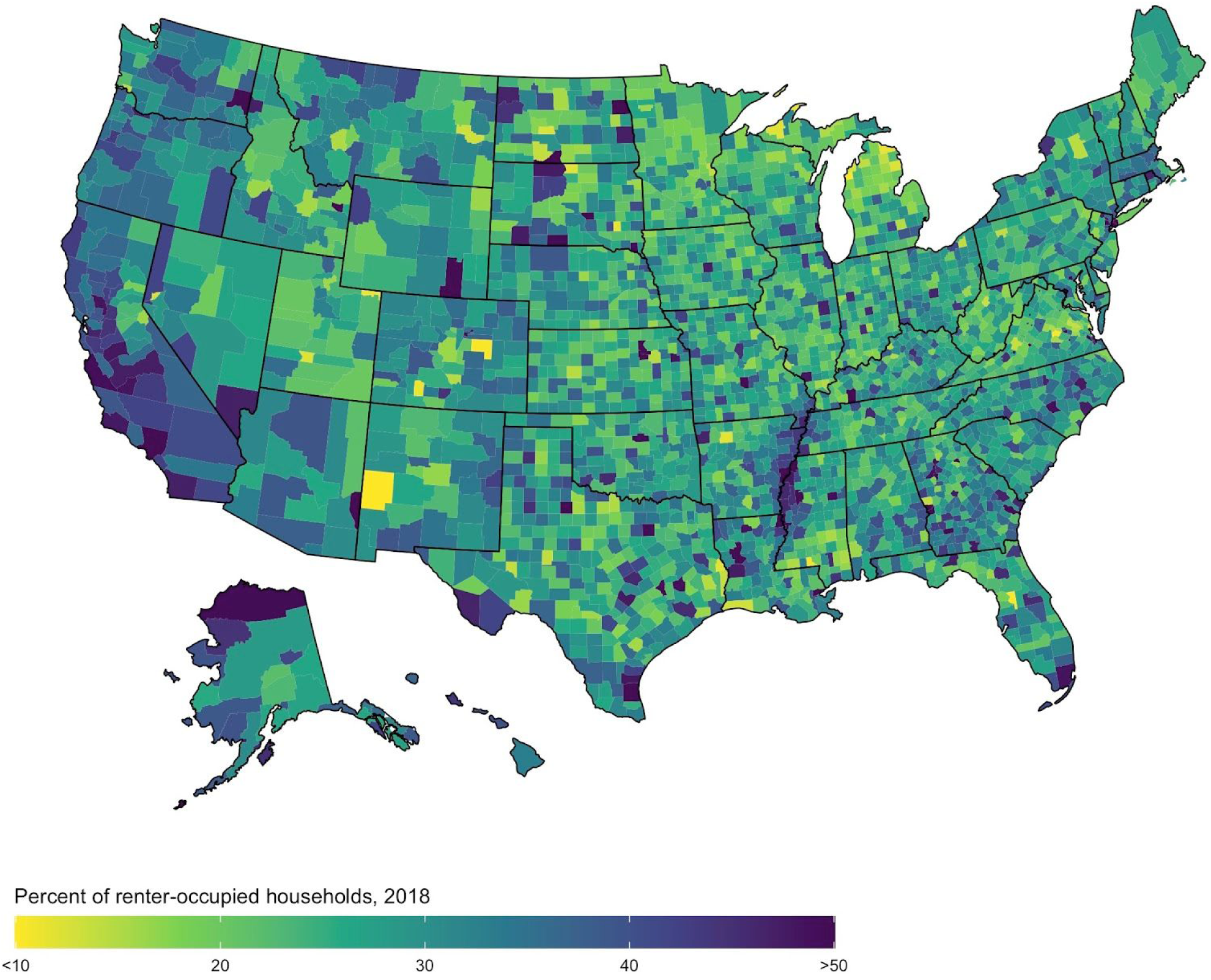
Percentage of renter-occupied households, 2018. Source: US Census Bureau’s American Community Survey 5-year, 2018 (variable S1101_C01_020E).

**Figure S15.**
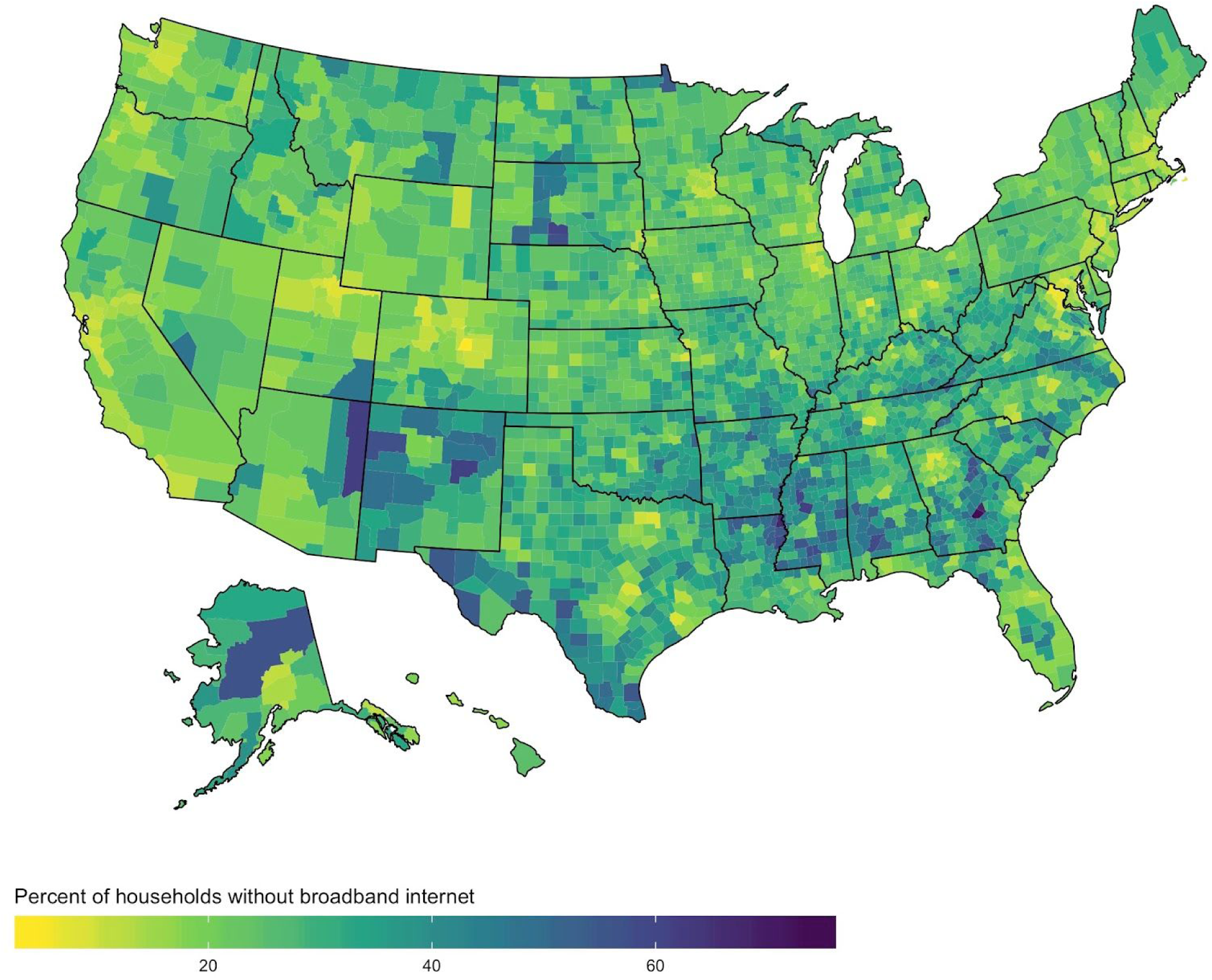
Percentage of households without broadband internet, 2018. Source: US Census Bureau’s American Community Survey 5-year, 2018 (100% -variable DP02_0152PE).

**Figure S16.**
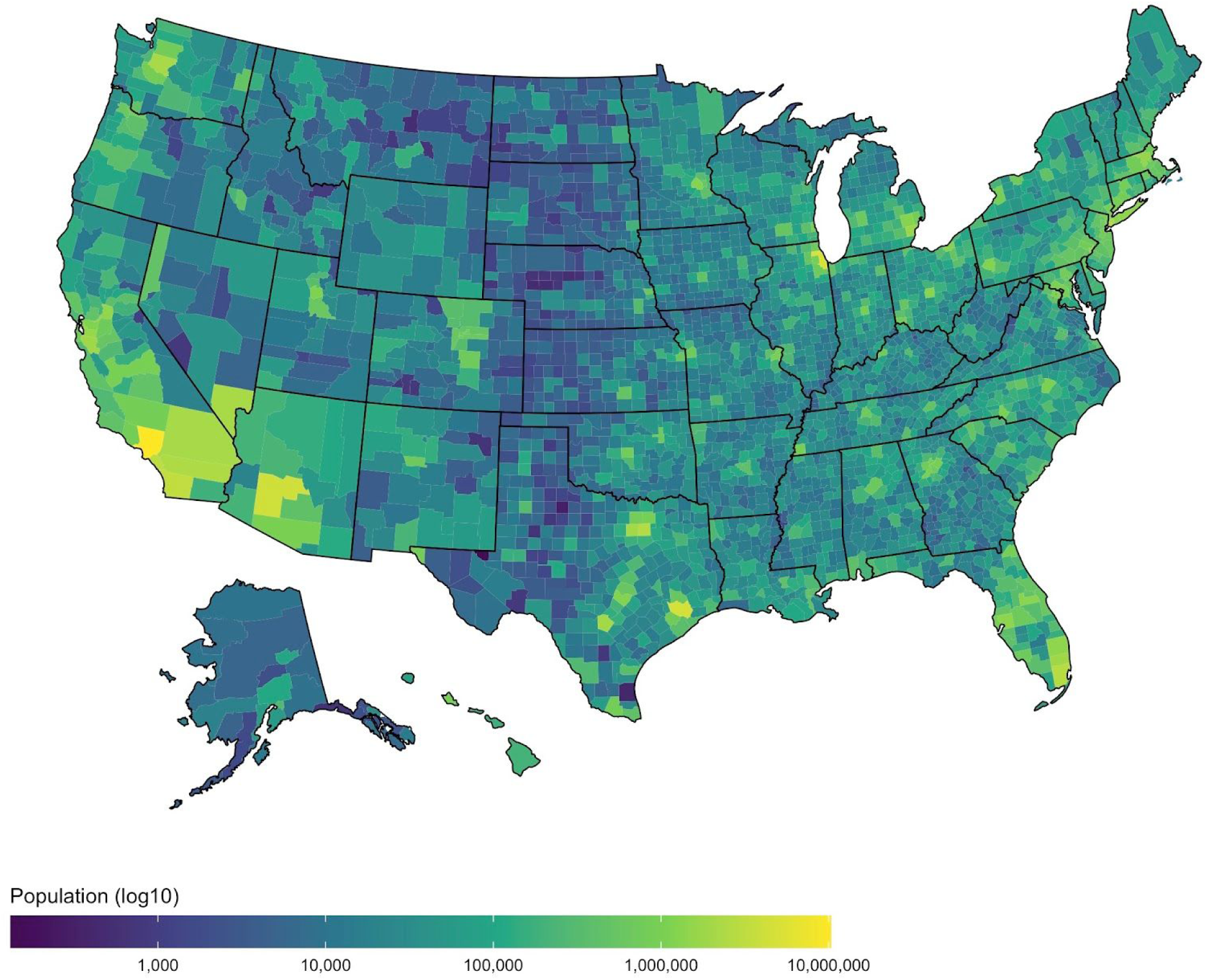
Population estimates (log 10), 2018. Source: National Center for Health Statistics (Bridged Race Estimates, Vintage 2018)

**Figure S17.**
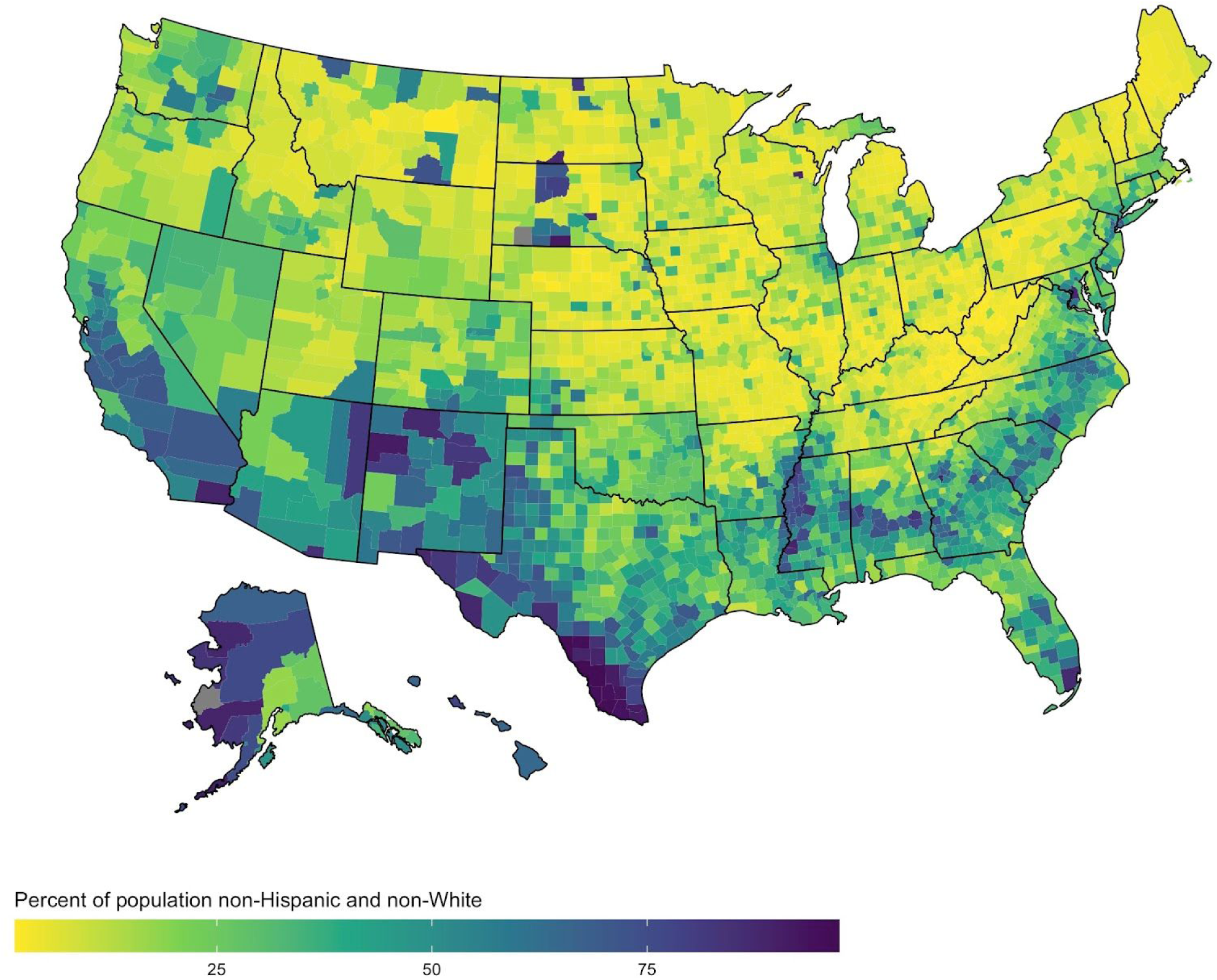
Percentage of population that is non-Hispanic and non-White, 2018. Source: National Center for Health Statistics (Bridged Race Estimates, Vintage 2018)

**Figure S18.**
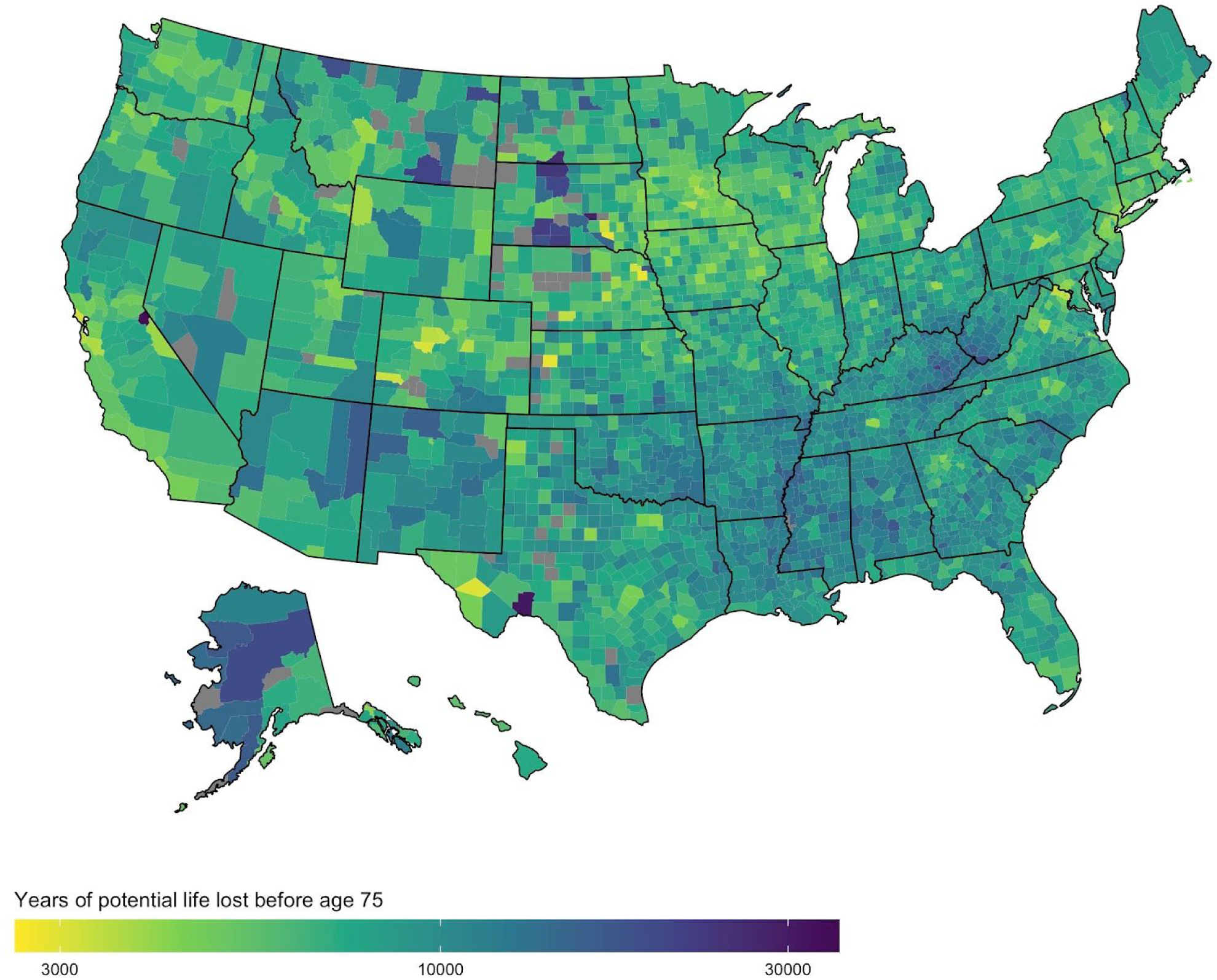
Age-adjusted years of potential life lost before age 75 per 100,000 population, 2017. Robert Wood Johnson Foundation County Health Rankings, 2019.

